# The McMaster Racialized Resident Mentorship Program Evaluation Protocol: Evaluating a racialized resident to racialized staff physician mentorship network on resident sense of belonging and medical training outcomes

**DOI:** 10.1101/2023.09.10.23295329

**Authors:** Anjali Menezes, Neha Arora, Curtis Sobchak, Marck Mercado, Madeline McDonald, Sandra Monteiro, Teresa Semalulu, Gina Agarwal, Suzanne Archie, the DARe Group Collaborative

## Abstract

The McMaster Racialized Resident Mentorship Program Evaluation will formally evaluate the effectiveness of a racialized resident mentorship network at increasing racialized residents’ sense of belonging to the medical training environment and reducing the racial attainment gap in medical careers. The program is composed of three phases. Phase 1 is an acceptability study of the collection of race0based data from all matriculating residents at McMaster University. Phases 2 and 3 will focus on family medicine residents. Phase 2 is a formal program evaluation of a mentorship network connecting matriculating racialized residents with racialized physician mentors, intended to run for 24 months and using repeat focus groups to explore the impacts of the program on residents’ sense of belonging. Phase 3 is a cross sectional study of graduating family medicine residents, examining associations between residents’ identities and attainment within residency training.

The program evaluation will involve formal mentor training, a mentor Community of Practice, and mentor and resident focus groups examining experiences within the program and sense of belonging to the Department of Family Medicine and their training sites. The program process inputs will include: the number of faculty who attend mentor training, and the number of racialized faculty mentors and racialized residents who register for the program; attendance at the Community of Practice; and attendance at focus groups of mentors and residents. An exit survey will assess the number of residents who participated in the program, the duration of participation, expected graduation time, number of mentor meetings. Short term outcomes will be measured at the phase 1 survey and at program enrollment and include: resident awareness of racialized mentors, and sense of support and sense of belonging. Long-term outcomes for the program will be assessed in phase 3, examining associations between resident social identities (including race) and family medicine residency training outcomes.

The project results will represent the first investigation of racial attainment in postgraduate medical training in Canada, with changes in residents’ sense of belonging and attainment during residency as indicators of Mentorship program effectiveness.

## INTRODUCTION

Racialized learners face unique challenges in their medical training, including overcoming interpersonal and structural racism from patients, educators, and peers.(1–5). While many new equity, diversity, and inclusion (EDI) initiatives striving to address overt racism and prejudices as well as systemic and structural racism affecting learners have been developed, in higher education and medical training, few have been evaluated and few indicators have been identified to measure program success. (6,7)

Differential attainment (DA) research studies racial disparities in medical learner and physician attainment – any measure of professional success, from exam performance to postgraduate employment rates, professor tenureship, and research funding allocation.(8–11) The United States (US) and United Kingdom (UK) have already taken bold steps to reduce racial differential attainment in medical careers.(10,11) Importantly, Canada has failed to recognize or address DA in medical training, as the Canadian medical profession has never collected race-based performance data, thus failing to identify baseline data essential for the evaluation of EDI programs striving to dismantle racism in our profession.(12)

Racialized patients need to see themselves reflected in their health care environment, through a workforce that exemplifies equity through its diverse membership, supportive and equitable training pathways, and professional self-awareness. The patient-physician relationship is central to the practice of medicine, a trusting relationship with patients and communities that is built on honesty and openness.(13) Physicians are symbolic representatives of the medical profession and health care system as a whole. We must be transparent in our self-assessments of the role racism plays in our own profession, and actively work to dismantle it with evidenced-based interventions and rigorous evaluations of their effectiveness.

Numerous studies exploring the experiences of racialized and minority trainees underrepresented in medicine have found a consistently expressed need for mentorship from mentors who are either racially congruent or from similar racial backgrounds (1,2,4,14,15). Mentorship is associated with increased career satisfaction, increased preparedness of junior faculty, increased research productivity and can increase learners SOB(5,7,14).

This project aims to institute a racialized resident and racialized staff physician mentorship network for all matriculating residents at McMaster University. We hypothesize that a mentorship network specifically designed and directed at racialized residents will increase their sense of belonging during their training and reduce the racial attainment gap in medical training outcomes.

The findings of this program evaluation will not only help promote equity in our physician workforce by increasing the retention and career satisfaction of racialized learners, but will also help to further our understanding of the mechanisms by which racialization impacts learning. Future applications of this foundational knowledge can be extended to other racialized populations, such as helping us to understand how the healthcare environment racializes patients and the factors that influence our sense of belonging and community.

### Program overview

Our protocol involves 3 phases of study: Phase 1 participants will be all matriculating first year (Postgraduate year 1, PGY1) residents; Phase 2 and Phase 3 participants will be family medicine residents, due to their shorter residency program length (two years) compared to other residency programs, and study funding and time constraints. An overview of each phase is provided below.

- **Phase 1:** An acceptability pilot study of a survey tool to collect race-based and other sociodemographic information from all matriculating residents that will serve as a baseline measurement of racialized learner attainment. Data collection of this part of our study is currently ongoing.
- **Phase 2:** A formal program evaluation of the effect of mentorship on racialized family medicine residents’ sense of belonging and obstacles encountered during their training. Primary data will be qualitative, collect through separate resident (mentee) and staff physician (mentor) focus groups, occurring three times a year until study completion.
- **Phase 3:** An exit survey collecting race-based and other sociodemographic information along with self-reported residency training outcomes, for all graduating family medicine residents, at the end of the second year from the implementation of Phase 1.

This program will apply theories of resiliency and established causes of DA from international frameworks to a racialized resident mentorship program and is expected to run for 2 years(5,16–18). We have chosen to focus on the construct of sense of belonging (SOB) as an fundamental contributor to academic success(5,7), with two studies specifically confirming its central role in the Canadian undergraduate medical training context (14,19).

Self-identified racialized residents will be matched to trained racialized staff mentors with the minimum requirement to meet at least three times per academic year. The program will use both quantitative (in the form of self-reported medical training outcomes at baseline in Phase 1, and at completion in Phase 3) and qualitative (in the form of separate mentor- and mentee-focus groups examining perceived impacts on sense of belonging) to evaluate the impact of racialized resident mentoring on residents’ sense of belonging and residency training outcomes.

## MATERIALS AND METHODS

### Objectives

#### Objectives for Phase 1

1. To describe the current diversity, racial and sociodemographic composition, and overall sense of belonging of the matriculating residents at McMaster University.
2. To obtain a baseline measurement of the current diversity of the resident cohort which can be used as a baseline for ongoing EDI interventions implemented at our university.
3. To explore resident respondents’ perceived acceptability of being asked to provide racial and sociodemographic information at matriculation.

#### Objectives of Phase 2

1. To pilot the implementation of a structured racialized mentorship network in postgraduate medical education.
2. To explore racialized family medicine residents’ views on the factors that contribute and inhibit the development of residents’ sense of belonging to their residency training community and overall training experiences.
3. To explore racialized staff mentors’ views on the factors that contribute and inhibit the development of racialized family medicine residents’ sense of belonging to their residency training community and overall training experiences.

#### Objectives for Phase 3

1. **The primary objective** will be to describe the associations between graduating family medicine residents’ racial sociodemographic identities, and residency training outcomes.
2. **The secondary objective** will be to describe the association between racialized family medicine residents’ participation in our mentorship program, overall sense of belonging, and residency training outcomes.
3. **The tertiary objective** will be the association of graduating family medicine residents participation broadly in any mentorship relationship (both formal and informal programs) and residency training outcomes.

### DETAILED DESCRIPTION OF PHASE 1

Acceptability study for the routine collection of race-based, sociodemographic, and performance data of matriculating postgraduate year 1 residents. A voluntary self-administered questionnaire will be used to collect baseline data and assess the interest of matriculating racialized residents in enrolling in a racialized mentorship program. As no routine data on the racial and ethnic make-up of family medicine residents is collected at this institution, this phase of the study will also allow the research group to assess the size and make-up of the racialized resident population. As this data is not routinely collected in the Canadian context, it will provide an overview of the perceived acceptability of the routine collection of this data to guide future advocacy for systematic data collection in this research field.

#### Participants

Phase 1 study participants will be all matriculating PGY1 residents at McMaster University beginning in July 2023.

#### Data Collection + Variables

Data will be collected using a self-report questionnaire emailed via REDCap, along with information about the pilot study to all incoming residents at McMaster University. The invitation to participate in the study and survey link will be emailed at the beginning of July 2023, with a reminder email sent once per month in August, and September 2023. The study information and consent form will include an explanation for the reason we are collecting race-based data, with a clear statement that participants’ information and individual racial identity disclosures will not be given to their residency program and will not affect their residency training progression. Additionally, the primary investigator (AM) will present an overview of the study at the residency orientation day in July.

The racial categories used for this pilot study will primarily follow the Canadian Institute for Health Information (CIHI) 2022 guidelines on the Use of Standards for Race-Based and Indigenous Identity Data Collection and Health Reporting in Canada (20). Adjustments were made to these proposed racial categories to ensure the dataset is comparable to other race-based data being collected or proposed to be collected by medical education bodies in Canada,(21,22), and were made following in depth discussions with the Differential Attainment REsearch Group Collaborative. We will take the additional steps of asking participants to self-identify as racialized or non-racialized. *Racialized* is a new term in the Canadian health context(23) and remains undefined. This additional data will provide information about how different races self-identify within this construct. We will then ask participants if they find it acceptable to be asked to provide their racial identity as a routine part of their employee onboarding.

We will invite racialized participants to state their interest in a racialized staff mentorship program. Those answering in the affirmative will be asked if they would prefer a staff mentor of the same racial background as themselves (with the clause that this could not be guaranteed but every effort would be made to help facilitate this). If racialized participants choose to participate in the mentorship program, they will be asked to provide their email to send mentorship information and phase 2 consent forms.

The survey will also collect data on gender identity, age, household income, Undergraduate GPA, and pre-medical school MCAT score, highest level of education achieved before medical school, current number of dependents, first language spoken, highest level of education achieved by parents, and family household income of parents. It will also ask residents to rate their sense of belonging in the medical training program (a metric designed to be comparable with national Canadian data from Statistics Canada (24), as well as their current access to mentorship support, on a 5-point Likert-like scale.

The final question of our survey provides residents with a free-text response box to disclose any concerns they may have about the standard collection of the information in our survey from medical trainees.

#### Consent

The participants were provided with a detailed letter of information for the study online, and each participant provided informed consent online through an electronically signed online consent form via our online survey tool: REDCap. Participants were provided with contact information to reach the study coordinator in case of any questions or concerns after reading the letter of information.

#### Plan for Analysis

Descriptive univariate statistics will be conducted on all the data, and the distributions examined. Two sample t-tests will be sued to compared mean GPA and overall MCAT scores between the racialized and non-racialized resident groups. This is the first study in Canada examining the racial identity of residents so estimations of the racial make-up of the resident cohort are difficult. Subgroup analyses will only occur if there are sufficient subgroup sizes. Chi-squared tests will be run to compare racial identity with self-reported acceptability of providing racial identity information. ANOVA will be run for racial identity and Likert-scale data. Qualitative description will be used to analyze any responses gathered from our free-text response option on the acceptability of our survey questions.

A logistics regression model will be used to examine the influence of all variables of interest on survey acceptability.

### DETAILED DESCRIPTION OF PHASE 2

#### Mentorship program objectives

1. To understand the impact of a racialized mentorship network on racialized family medicine resident’s sense of belonging
2. To evaluate the efficacy and value of mentorship programs on racialized family medicine resident residency training outcomes.

#### PROGRAM LOGIC MODEL

The program will consist of trained racialized family physicians being voluntarily matched to incoming racialized family medicine residents with a requirement of meeting at least three times a year to discuss their career and their experiences in medicine. Our program logic model is described in more detail in the supplementary materials.

The proposed intervention, which will be implemented for all incoming first year residents, will involve the following steps:

##### Phase 2a: Training Faculty Mentors

- An email will be sent to all Faculty in the Faculty of Health Sciences at McMaster, seeking volunteers to join the racialized staff mentorship network and advising non-racialized faculty of the program.
- All faculty who register to be mentors will be asked to complete a mentor registration form that will ask for their racial identity, their speciality, and their preference to be matched based on their race, or specialty.
- Mentors will be expected to attend 4-hour training workshop which will review differential attainment, and literature on its theorized causes. The workshop will outline techniques to support racialized learners, including culturally aware mentorship, and how to build resilience and enhance the steering effect and self-complexity of learners who experience discrimination during their training.

##### Phase 2b: Advertising and Running the Mentorship Network

- As part of the collection of baseline characteristics described in Phase 1 of the program, residents had the opportunity to self-identify for the mentorship network. These residents will be contacted about this program, provided with a formal consent letter and asked to register for the program.
- Residents who register with the service will be matched to a racialized staff mentor, and a first meeting will be arranged by the mentor. While no specific outline for the first mentorship meeting will be mandated, each mentoring dyad will be provided with a “Mentee Goals & Objectives” form to help guide their relationship. We have followed the conceptualization of mentorship in the graduate medical education setting of DeTurk, Kaza, and Pellegrino which outlines the role modelling, educational, wellness and counselling dimensions of mentoring relationships (25). Mentors will be encouraged to adjust their approaches depending on the individual expressed needs of their mentee. We have chosen this unstructured format in light of the widely varying possible foci for mentorship programs(1) in the context of our objectives and hypothesis that any mentoring relationship will impact perceived sense of belonging and thus impact residency training outcomes.
- The minimum meeting requirement will be once every 4 months, at which point, the “Mentee Goals & Objectives” form will be reviewed. Mentees and mentors will be advised that there is no maximum number of mentoring meetings they can have.

##### Phase 2c: Assessing and Monitoring the Mentorship Program

- Advertising effectiveness information will be collected on the number of people who opened the advertising email, the number of faculty and residents who enquired about it, and the total number of faculty and residents who signed up for the program.
- Additionally, information on the number of faculty who signed up for the workshop, along with general feedback for the workshop through a self-administered survey exploring comfort levels with discussing race; discrimination; and mentoring trainees, will be conducted, using pre- and post-workshop ratings on 5-point Likert-like scales, and the option to provide qualitive feedback in free-text response boxes.
- Before their first mentorship meeting takes place at the beginning of residency, residents will be invited to a 45-minute focus group that will explore residents’ perceptions of:

a. Their sense of belonging to their residency program community
b. Their overall views of the goals, possible impacts, and conceptualization of mentorship.
c. Experiences of inclusion and exclusion during their mentoring training to date
d. The factors their feel may influence their ability to *attain* during medical training.
- Mentors will also be invited to a 45-minute focus group before they meet their mentee which will explore the same topics, with the adjustment that point (c) and (d) as outlined about will be phrased to explore mentors perceptions of their mentees’ experiences.
- Monitoring focus groups for residents’ will be run again in January and May of year 1, and July, December, and May of year 2. These focus groups will explore residents experiences of belonging and exclusion during their training and their relationships with their mentors.
- A faculty focus group will also be run at these points which will examine faculty perceptions of how they feel their mentoring role is going, general themes emerging from their mentees regarding their stressors, sense of belonging, and any experiences of discrimination their mentees faced and how these were navigated.
- The program will also run a Community of Practice for faculty mentors three times a year, to support mentors, problem solve support techniques, share best practices, and to monitor for any trends or concerns in resident wellbeing and safety.

#### RESEARCH PARTICIPANTS

Participants will be all incoming, Canadian medical school graduates of the McMaster Family Medicine Residency Program of July 2023, and their racialized faculty mentors. International medical graduates will be excluded as racialization is context specific.(12)

#### Consent

Focus group participants will be provided with a printed Phase 2 Letter of Information which they will sign before entering the focus group room. Informed consent process will be administered by the study coordinator. In addition, each focus group interview will begin with a statement reminding participants of the letter, and neither the focus group nor recording of the focus group interview will begin until each participant verbally re-affirms their consent.

#### RESEARCH OUTCOMES

Broadly, the program evaluation will involve the collection of both descriptive and qualitative datasets, outlining:

- Organizational outputs: emails, announcements, survey links to residents and faculty
- Engagement with the program: faculty registration and attendance rates for the program and mentor training; attendance rates at faculty Community of Practice meetings and both faculty and resident focus groups; the number of mentoring meetings attended
- Qualitative data on the impacts of the program on the resident sense of belonging and comfort and confidence in navigating any instances of discrimination or marginalization in their postgraduate training. Our focus group interview guide is provided in the Supplementary materials and provides further details.
- Qualitative data on faculty mentors’ preconceptions about mentorship, sense of belonging and experiences with Community of Practice. Our focus group interview guide is provided in the Supplementary materials and provides further details.

#### Stopping criteria for qualitative data

Our study sample will be recruited through convenience sampling for our focus groups from the family medicine mentorship dyads included in our program. Due to funding and time constraints, we will only be able to conduct three sets of focus groups per year, with these pre-specified timings planned to correlate with natural transitions during residency training.

#### Data analysis plan

All quantitative data collected for Phase 2 will be analysed using descriptive univariate statistics.

The qualitative data will be analyses using interpretive phenomenological analysis (IPA) approach through the lens of critical race theory (CRT). This approach was chosen for two reasons. As a theoretical perspective, CRT posits the ordinary and covert everyday functions of racism within structures and institutions.(26–28) As we aim to explore how mentorship can interact with the many forms of racism that shape the identities of racialized residents, to influence the everyday lived and broad construct of our sense of belonging, CRT allows for this.(27,28) We have chosen to use IPA as it seeks to understand the meanings participants attach to their lived experiences and our objectives explore how individual experiences in residency training and within mentor dyads influence our sense of belonging.(29) We will adopt an approach as described by Williams et al (2022) in applying both CRT and IPA to data analysis(28).

The research team is purposefully almost exclusively racialized with a diversity in the racial identities of our investigators. We specifically include Black, Indigenous, and Filipino team members as representatives of underrepresented minorities in medicine in our specific context (30,31). Our whole research team will be involved in analysing data as this diversity of our analytic team will help to provide the racial context needed for understanding the nuances of the interpretations of racism.

We will circle back to our focus group participants for member checking, and will develop themes through consensus building among team members. Reflective logs of how our own experiences with racism have resurfaces throughout this process will be conducted regularly.

### DETAILED DESCRIPTION OF PHASE 3

This phase of the project will be a cross-sectional study at the end of our intervention, looking for associations between resident racial identity, participation in the program, and several measures of “attainment” during residency training.

#### PARTICIPANTS

Participants will be all graduating Family Medicine residents at McMaster University in 2025

#### Methods

Data will be collected using a self-report questionnaire emailed, along with information about the study to all graduating family medicine residents at McMaster University in 2025. The invitation to participate in the study and survey link will be emailed in April 2025, with a reminder email sent once per month in May, June, and July 2025. The study information and consent form will include an explanation for the reason we are collecting race-based data, with a clear statement that participants’ information and individual racial identity disclosures will not be given to their residency program and will not affect their residency training progression.

The survey will be sent to all graduating family medicine residents, as datasets from residents who did not participate in the mentorship program will be used as our comparison group. The collection of race-based data along with postgraduate training outcomes will represent the first DA research in postgraduate medical training in Canada.

##### Race

The racial categories used for this pilot study will primarily follow the Canadian Institute for Health Information (CIHI) 2022 guidelines on the Use of Standards for Race-Based and Indigenous Identity Data Collection and Health Reporting in Canada statement (20). Adjustments were made to these proposed racial categories to ensure the dataset is comparable to other race-based data collected or proposed to be collected by medical education bodies in Canada (21,22), and were made following in depth discussions with the Differential Attainment REsearch Group Collaborative.

##### Other social identity and socioeconomic class data

The survey will also collect data on gender identity, age, household income, highest level of education achieved before medical school, current number of dependants, first language spoken, highest level of education achieved by parents, and family household income of parents.

##### Mentorship Program Participation

The Phase 3 survey will ask residents if they ever had a mentor during their residency training, and if so, a free text response option will ask them to provide the name (if it was through a formal mentorship program).

##### Likert-like scale questions

Similarly to Phase 1, we will ask residents to rate their agreements with two statements: that they have a strong sense of community to their medical training program; and that they have had mentorship support during their medical training.

##### Residency Program Attainment

The racial attainment gap in standardized and high stakes testing at all levels of education, including medical training are well established (32,33). We have chosen not to further stigmatize racialized medical learners by comparing certification exam scores. Likewise, the College of Family Physicians of Canada’s (CFPC) Triple C Framework outlines the goals of residency training in producing competent family physicians (34). It outlines the inadequacies of assessing competency through multiple choice or objective structed clinical examinations. Instead, residents are assessed through recurrent reviews of the cumulative evidence of learners demonstrated competency in multiple settings (34). The metrics collected to assess residents’ attainment during training have thus been informed by the Triple C Framework, as implemented at McMaster University, and the PI’s (AM) role as the anti-racism advisory on the central competency committee for the family medicine residency program under study. Our stakeholders, the residency program director and the dean of postgraduate medical education have additional reviewed our chosen metrics.

Are survey will ask residents to self report:

- Any leaves of absences from the program, and their length.
- If they passed the first sitting of their CFPC Certification Examination in Family Medicine, a combination of passing both the Short answer management problems(SAMPs) and simulated office orals (SOOs) component of the examination (35).
- Individual pass rates of the SAMP and SOO components respectively
- If the resident completed any remediation during their training self-reported by resident.
- If the Resident completed any enhanced education plans during their training self-reported by resident.

#### Consent

The participants will be provided with a detailed letter of information for the study online, and each participant will provide informed consent online through an electronically signed online consent form via our online survey tool: REDCap. Participants will be provided with contact information to reach the study coordinator in case of any questions or concerns after reading the letter of information.

#### Plan for Analysis

Descriptive univariate statistics will be conducted on all the data, and the distributions examined. This is the first study in Canada examining the racial identity of family medicine residents so estimations of the racial make-up of the resident cohort are difficult.

Subgroup analyses will only occur if there are sufficient subgroup sizes. If we proceed to subgroup analysis, where the responses will be evaluated based on the specific minority group, we will undertake community engagement or inclusion of other researchers who will focus on this direction for this research. We will submit an amendment to the ethics board to add this step.

Chi-squared tests will be used to compare the pass rates of first-time CFPC Certification in Family Medicine exams of racialized and non-racialized residents. A logistic regression model will be used to examine the influence of all variables of interest on CFPC exam pass rates. A two-sample t-test will be used to compare the mean CFPC exam score of the racialized with the non-racialized group. Correlation coefficients will be calculated for the influence of ordinal and continuous data on exam scores. One Way ANOVA tests will be used to examine the influence of the categorical data on exam scores.

## ETHICS

As this is the first planned collection of race-based performance data in medical careers in Canada. In accordance with the CIHI Guidance, our research team is broad and specifically includes Black and Indigenous team members who have actively contributed to the development of the project protocol and have access to our data (20). We also included Filipino members, who have been identified as underrepresented in the Canadian medical profession (30,31). We have also discussed our project plan with the Family Medicine Residency Program Director and the Associate Dean of Equity and Inclusion of the Faculty of Health Sciences of McMaster University.

### Indigenous Data Sovereignty

Working towards reconciliation, we recognize Indigenous peoples’ right to maintain, control, and protect their knowledge, culture, and intellectual property. Following The Tri-Council Policy Statement 2’s regulations on research ethics in Canada (36), we have additionally included our local Indigenous community in the conceptualization of our protocol and will continue to seek their input throughout our research process, at regular meetings. We seek to incorporate any and all of their recommendations to our research process at any stage in our project, and will submit amendments to our ethics board approval as needed.

### Ethical Approval

Ethics approval has been received from the Hamilton Integrated Research Ethics Boards (Project ID 15807). Approval was also obtained from the Research in Residency Committee of the Family Medicine Residency Program.

## STAKEHOLDER ENGAGEMENT

As this study is first of its kind and some of these phases include inter-departmental collaborations with the McMaster Faculty of Health Sciences, we have included the following stakeholders to this study as they are people who have an interest in our research project or are affected by its outcomes. We plan to periodically share the interpretations and study findings with our stakeholders.

Our stakeholders are as follows:

- Family Medicine Residency Program Director, McMaster University
- Chair, Indigenous Health, UGME, and our study liaison with the Indigenous Health Learning Lodge, McMaster University
- Dean, Post-graduate Medical Education, McMaster University

All the data shared will either be anonymous or de-identified and will be consolidated in terms of descriptive statistics or thematically for qualitative data.

## PLAN FOR DISSEMINATION

We plan to prepare three separate publications of the results of our project (one for each phase) to be submitted to high impact medical education research journals. We plan to present findings of each phase of our project at the International Congress on Academic Medicine. The research findings will broadly influence the ongoing training provided by the DARe Group Collaborative to faculties across North America on proven evidence-based interventions to address racial DA. We plan to eventually extend our work to postgraduate medical training programs across Canada.

## STRENGTHS AND LIMITATIONS

Our study has a number of strengths. It will be the first study exploring residents’ acceptability of the collection of race-based data at training matriculation. Our survey tool has been developed with the input of the DARe Group Collaborative – a pan-Canadian national network of racialized academics aiming to measure and work to eliminate the racial attainment gap in health professional training and career outcomes. This national alignment is important, as the collection of race-based data is relatively new in the Canadian medical context and, from the diverse work contexts of our study team, there are many different tools in varying process of development and implementation to collect this data across the country. Working towards national partnerships is essential, for the alignment of metrics that are comparable and thus paint a more fulsome picture of the Canadian context.

Mentorship programs are rarely evaluated comprehensively with quantitative measurements (1), let alone the both qualitative and quantitative metrics in our mixed-methods approach. Our evaluation approach is additionally novel as it seeks to directly assess the impact of mentorship on a widely recognized factor influencing racial differentials in attainment. In choosing this approach, our findings will make in important contributions to antiracism research in two ways. First, as we are focussing on changes in SOB, they will be able to contribute to the limited literature on the extent to which our sense of belonging is impacted by structured organizational EDI interventions. And secondly, in choosing to incorporate measures of attainment in our study, we have set a standard for the evaluations of EDI interventions that seek to dismantle racism in our profession.

We anticipate two limitations to our study. Our surveys will be optional and as we know that survey response rates are often low, in the context of a study at a single institution, and within a single training program (for Phase 3), our projected sample size will be small. Secondly, while we have created an exclusively racialized mentorship network, our probable small sample size will mean making all mentorship dyads racially congruent unfeasible.

## CURRENT STATUS AND PROJECTED STUDY TIMELINE

Phase 1 of the study is funded by the Racialized Community Leadership Grant in Family Medicine from the Foundation for Advancing Family Medicine of the College of Family Physicians of Canada. Phase 2 and Phase 3 of the study are funded by the McMaster Academic Family Medicine Associates.

Phase 1 of our study is underway, currently in the process of data collection. Phase 2 of our study is underway, currently recruiting and training staff mentors, but data collection has not begun. Phase 3 of our study has not been initiated to date.

## AUTHOR CONTRIBUTIONS

### Conceptualization

Anjali Menezes, Curtis Sobchak, Teresa Semalulu, Marck Mercado, Madeline McDonald, Sandra Monteiro, Gina Agarwal, Suzanne Archie.

### Funding Acquisition

Anjali Menezes, Gina Agarwal.

### Methodology

Anjali Menezes, Sandra Monteiro, Gina Agarwal, Suzanne Archie.

### Writing - original draft

Anjali Menezes, Neha Arora.

### Writing – review and editing

Anjali Menezes, Neha Arora, Gina Agarwal.

## SUPPORTING INFORMATION

S1. Phase 1 survey tool

S2. Phase 2 Program Causal Model and Theory of Action

S3. Phase 3 Interview Guide

## Data Availability

No datasets were generated or analysed during the current study. All relevant data from this study will be made available upon study completion.

